# Enhanced versus standard outpatient care of asthmatic children in Malawi: study protocol for a pilot randomised controlled trial

**DOI:** 10.1101/2020.08.12.20173401

**Authors:** Sarah Rylance, Joseph Phiri, Jonathan Grigg, Chris Jewell, Kondwani Jambo, Kevin Mortimer

## Abstract

**Background:** Asthma is the most common chronic disease in childhood and is a growing global concern. However, there are limited data regarding long-term asthma management in low-income countries (LIC), particularly for children.

This study will assess the feasibility of recruitment and retention, the acceptability of an intervention and data collection methods, and baseline levels of asthma control and exacerbation rates in our target population of Malawian asthmatic children. Furthermore, we plan to; evaluate the effect of a package of enhanced asthma care, appropriate for a LIC, over a 3-month period; to describe clinical and airway inflammatory phenotypes; to identify clinical and laboratory features which might predict response to treatment in this population.

**Methods:** We will recruit 120 children aged 6-15 years, attending outpatient asthma follow-up at a tertiary government hospital in Malawi. Participants will be randomised, stratified by level of asthma control (Childhood Asthma Control Test (C-ACT) ≤19 or ≥20), to receive standard care in the hospital clinic, or an enhanced care package comprising; detailed clinical assessment (including pre- and post-bronchodilator spirometry, exercise challenge, exhaled nitric oxide (FeNO) measurement and sputum induction), optimisation of inhaled asthma treatment, and asthma education delivered by non-healthcare workers. Feasibility outcomes will include recruitment and retention rates, data completeness for study procedures, and baseline asthma control and exacerbation rates. The primary clinical outcome is asthma control, measured by C-ACT at 3 months, with adjustment for baseline measurement and intervention as covariates in a regression model. Secondary clinical outcomes at 3 months include; asthma exacerbations (requiring emergency health care use, treatment with oral corticosteroids or hospital admission), school absence, lung function and FeNO levels. Participants will be described by clinical and inflammatory phenotypes, with the latter categorised as eosinophilic or non-eosinophilic based on sputum cytology results and FeNO levels.

**Discussion:** It is important to assess whether global treatment guidelines for long-term asthma management are appropriate for low-income settings. This study will provide key feasibility data, including baseline clinical characteristics of asthmatic Malawian children, to inform assessment of an asthma care package in a low-income setting, which includes task-shifting to non-healthcare workers.

**Trial registration:** Pan African Clinical Trials Registry: PACTR201807211617031. Registered 09/07/18, https://pactr.samrc.ac.za/TrialDisplay.aspx?TrialID=3468

## Background

Asthma is the most common chronic condition in childhood and an emerging health problem in Africa (1). However, the burden of chronic respiratory symptoms in sub-Saharan Africa is poorly described. Limited data from urban populations estimate the prevalence of “current wheeze” at 14-20% among older children, and 5-13% in younger children, with severe asthma symptoms in over half of those reporting wheeze (2, 3). In Queen Elizabeth Central Hospital (QECH), a tertiary government hospital in Malawi, children frequently attend emergency care with acute wheeze, with severe cases admitted for nebulised and intravenous bronchodilators. Children with asthma are followed-up in a general paediatric clinic, alongside patients with a wide variety of other medical conditions. Due to the busy, unstructured nature of this clinic, it is challenging to deliver good quality long term management for specific chronic conditions.

The long-term goals of asthma management are to achieve good symptom control and minimize future risk of adverse outcomes (exacerbations, fixed airflow limitation and side-effects of treatment) (4). Current global treatment guidelines, focusing on inhaled corticosteroids (ICS), are extrapolated from high-income settings (4). These guidelines are appropriate where steroid-responsive, allergen-driven, eosinophilic asthma predominates. If the pathophysiology underlying wheeze in LIC is different, current recommended treatments may be ineffective or even harmful, particularly in the context of a high burden of tuberculosis (TB) and other opportunistic infections (5).

There are limited data on long term asthma outcomes for patients from low-income African settings; published studies relate to adult populations, with no data from paediatric patient groups. Among a Ugandan cohort during 1-year of follow-up, 60% experienced at least one exacerbation and one third experienced three or more.(6) Asthmatic patients attending teaching hospital clinics in Nigeria and Uganda reported high rates of uncontrolled asthma symptoms (defined as Asthma Control Test score ≤19); 57% and 83%, respectively.(7, 8) The burden of asthma symptoms and exacerbations is poorly defined in the paediatric population, and it is unclear whether asthmatic children in low-income settings respond to globally advocated treatment.

A search of the ISRCTN Register of Controlled Trials database and the WHO Pan African Clinical Trial Registry for asthma trials in Africa identified a total of 12 trials, excluding this one, all from middle-income countries; South Africa (5 trials), Egypt (6 trials), and Kenya (1 trial).

The piloted intervention will include the components of global treatment guidelines that are feasible and potentially scalable in Malawi, at the present time. Due to the lack of asthma research in paediatric populations from LIC, it is necessary to assess trial feasibility, including recruitment and retention rates, acceptability of the intervention and data collection methods, and baseline levels of asthma control and exacerbation rates in our target population. Furthermore, the aims of this study are to; evaluate the effect of a package of enhanced asthma care, appropriate for a LIC, over a 3-month period; to describe clinical and airway inflammatory phenotypes of asthmatic children in Malawi; to identify clinical and laboratory features which might predict response to treatment.

## Methods/design

### Study design

This is a pilot randomised controlled trial of standard vs enhanced asthma care, in a cohort of children attending outpatient follow-up at a tertiary, government hospital in southern Malawi. Additionally, we will use clinical and laboratory methods to describe the phenotypes of participants and identify factors predictive of poor asthma control.

### Study intervention

Participants will be randomly assigned to receive standard or enhanced care, with stratification by baseline level of asthma control, defined by Childhood Asthma Control Test (C-ACT) score; good control if ≥20 or poor control if ≤19.(9)

#### Standard care

Children will continue to receive care in the general paediatric clinic at QECH. Short acting β_2_ agonists (SABA) (e.g. salbutamol) and inhaled corticosteroid (ICS) (e.g. beclomethasone diproprionate (HFA-BDP)) inhalers are included in the Malawi Standard Treatment Guidelines, and usually available, free of charge, in the hospital pharmacy. Dose adjustment of inhaled treatment, patient education and follow-up interval are at the discretion of the reviewing clinician (a registrar or consultant Paediatrician).

#### Enhanced care

The enhanced care intervention will have 3 components; detailed clinical assessment (including pre- and post-bronchodilator spirometry and exercise challenge), optimisation of inhaled asthma treatment, and asthma education. Fieldworkers (non-healthcare workers) will deliver a 1-hour asthma education session to the child and carer, including discussion of a personalised asthma action plan, with oversight by the study clinician or nurse. Before study initiation, fieldworkers will complete a structured training programme, with accompanying training manual, and will complete an assessment of their knowledge and skills. Each asthma education session will follow a structured approach, with checklist, to ensure intra- and inter-fieldworker consistency.

Following a detailed clinical assessment, ICS treatment will be escalated to 200μg twice daily of HFA-BDP, via a home-made bottle spacer, for those children identified as having poor asthma control, in line with global asthma guidelines (4). This subgroup of patients will be reviewed at 6-weeks, for repeated asthma education and to assess their response to treatment and treatment adherence. Those with good control will continue their existing dose of ICS. All children will receive inhaled SABA as needed, via a home-made bottle spacer.

### Study population

Participants will be recruited from the General Paediatric outpatient clinic at Queen Elizabeth Central Hospital (QECH), in Blantyre, Malawi. This clinic receives referrals from primary health care centres, the QECH Paediatric Emergency Department, and following discharge from the paediatric wards at QECH.

### Eligibility criteria

All assenting children aged 6-15 years attending the QECH paediatric general clinic with doctor-diagnosed “asthma” will be eligible for inclusion. Children receiving TB treatment or those with symptoms suggestive of TB (fever, night sweats, haemoptysis, weight loss) will be referred to the local paediatric TB clinic and excluded from the study.

The initial study visit will be postponed for patients who are acutely unwell, or currently receiving oral corticosteroids, or with a recent respiratory infection or asthma exacerbation (within past 4-weeks). Children with contraindications to spirometry (chest or abdominal pain, haemoptysis) will be excluded from spirometry, sputum induction and exercise challenge, but may participate in all other aspects of the study.

### Study procedures

Following written informed consent and assent, fieldworkers will administer an electronic questionnaire, including questions on; asthma symptoms, exacerbations, treatment and potential contributing factors. Children will be stratified as having good (C-ACT ≥20) or poor (C-ACT ≤19) control and randomised to receive standard or enhanced care. Random number sequences will be computer-generated, with variable, random length, permuted blocks. Separate sequences will be generated for those with good or poor control, with allocation derived from a concealed file which is accessible only during electronic questionnaire completion. Participants allocated to enhanced care will attend for a study visit shortly after recruitment, including; clinical examination, anthropometry, exhaled nitric oxide (FeNO), pre- and post-bronchodilator spirometry, exercise challenge and sputum induction, with a further visit at 6-weeks if they have poor control (C-ACT ≤19 or FEV_1_/FVC below the lower limit of normal) at the study visit. All participants will be reviewed after 3-months, including assessment of asthma symptoms, exacerbations, treatment, FeNO, and pre- and post-bronchodilator spirometry.

All study procedures will be performed according to standardised operating procedures, with regular quality control monitoring by the study clinician. Spirometry will be conducted by a trained and experienced technician, using an Easy On-PC spirometer (ndd Medical Technologies), according to American Thoracic Society/European Respiratory Society (ATS/ERS) guidelines (10). Lung function will be measured before and after a 400μg dose of Salbutamol administered by metered-dose inhaler via a Volumatic spacer. Lung function parameters; forced expiratory volume in one second (FEV_1_), forced vital capacity (FVC), FEV_1_/FVC ratio and bronchodilator responsiveness, will be interpreted with reference to the African American Global Lung Initiative 2012 equations.(11) Carboxyhaemoglobin will be measured non-invasively using a Rad-57 Pulse CO-Oximeter (Masimo Corporation). FeNO will be measure using an NObreath machine (Bedford Scientific Ltd.), as per ATS/ERS guidelines. (12) During the exercise challenge, participants will run, with maximum effort for a period of six minutes, with lung function (FEV_1_) monitored at five and ten minutes post-exercise. After recovery from exercise challenge, sputum will be collected, following sequential 5-minute nebulisation (Schill Multisonic Infracontrol) periods of 4.5% saline, to a maximum of 15 minutes.

Following the initial 3-month trial period, those children allocated to the standard care arm will be given the option to enter the enhanced care arm, for a further 3-month (post-trial) period. All participants will return to routine asthma care in the general paediatric clinic at QECH following study completion.

Procedures undertaken during the study visits are summarised in Figure 1.

**Figure 1.**
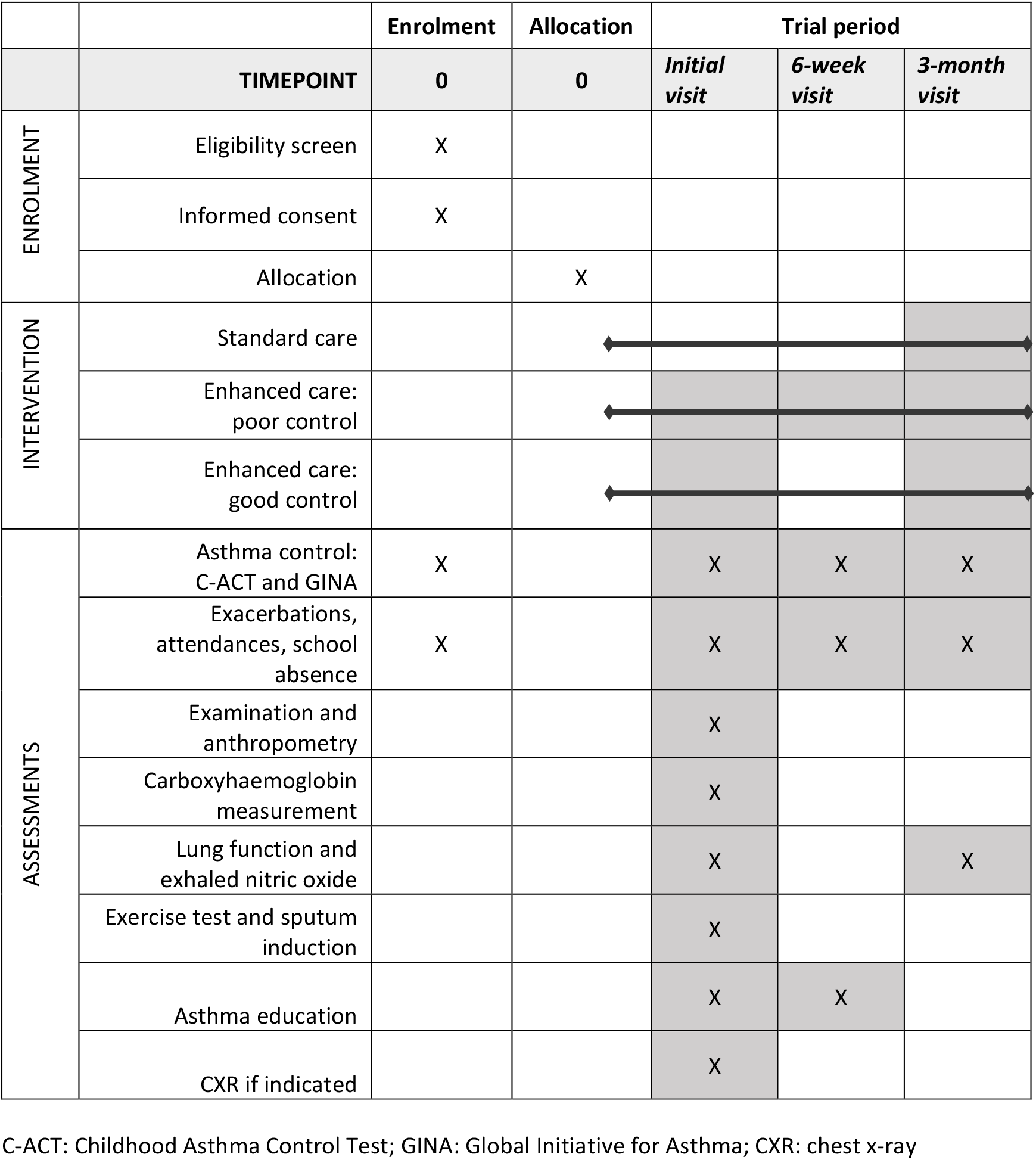
Schedule of trial procedures; enrolment, intervention, and assessments.

### Study risks

There is a small risk that study procedures (exercise challenge and sputum induction) may trigger asthma symptoms. Participants will be closely monitored during all study procedures by experienced medical staff, with access to inhaled and nebulised medications if required. A Data and Safety Monitoring Board (DSMB) will safeguard the welfare of trial participants: all serious adverse events will be reported to the DSMB within 72-hours of notification. Families are requested to contact the study team if their child is admitted to hospital, during the study period.

### Outcome measures

Feasibility outcomes will include recruitment and retention rates, data completeness for each study procedure, and baseline asthma control levels and exacerbation rates.

The primary clinical outcome is a composite score of clinical control, measured by C-ACT at 3-months. The C-ACT, a 7-item validated questionnaire, has been translated into Chichewa according to linguistic validation guidelines.(13) Secondary clinical outcomes are listed in table 1.

**Table 1.** Trial secondary clinical outcomes

|  | Timepoint |
| --- | --- |
| Exacerbations requiring hospitalization | 3 months |
| Exacerbations requiring emergency (unscheduled) health care use | 3 months |
| Exacerbations requiring systemic (oral) corticosteroid use for $\geq 3$ days | 3 months |
| Days absent from school | 3 months |
| Lung function (pre- and post-bronchodilator FEV<sub>1</sub>) | 3 months |
| Fractional concentration of exhaled nitric oxide (FeNO) | 3 months |

FEV<sub>1</sub>: forced expiratory volume in 1 second

### Laboratory procedures

Sputum samples will be transported on ice for immediate processing. Extracted mucus plugs will be homogenized with dithiothreitol, filtered, and prepared for further assessment by direct light microscopy and flow cytometry.(14) Both methods will assess the proportion of eosinophils and neutrophils, as a percentage of the total white cell count.

### Data collection and management

Electronic questionnaires will be completed using KoBo Toolbox software. Data will be backed up to the Malawi-Liverpool-Wellcome Trust server daily and reviewed regularly according to standard operating procedures to assess for inconsistent or missing data. All electronic data will be anonymised, except for the participant tracking form, which will be stored with double password protection. All paper documentation will be stored in a secure filing cabinet.

## Data analysis

The cohort will be described, in terms of proportions of patients demonstrating specific clinical and airway inflammatory phenotypes. Clinical phenotypes will be described by; atopic features, age at asthma onset, asthma triggers (including exercise), lung function, bronchial hyperresponsiveness (15). Airway inflammation will be categorised as eosinophilic or non-eosinophilic based on induced sputum cytology (≥2.5% vs <2.5% eosinophils/total sputum cell count respectively), and on the basis of FeNO results (low <20 ppb, intermediate 20-35 ppb, high >35 ppb)(16).

Feasibility outcomes will be reported descriptively and narratively. The primary clinical outcome will be reported as a scalar variable, defined by C-ACT score. The difference in mean C-ACT score between the intervention and control arms will be assessed using two-tailed Student’s t-test. To measure change in C-ACT over time, final measurements will be compared, with adjustment for baseline measurement and intervention, as covariates in a regression model. Data will be analysed by intention-to-treat. Per-protocol analysis will also be performed. The contribution of pre-specified explanatory variables; sputum eosinophilia, FeNO level and treatment adherence, will be assessed using multivariable linear regression using paired C-ACT measurements from participants (at the start and end of the enhanced care period).

### Sample size

Due to lack of previous data from comparable paediatric populations in Africa, we have designed the study based on several carefully considered assumptions; we have estimated population values using data from an adult cohort in Nigeria, which reported a mean ACT of 17.0 (standard deviation (SD) 5.3) and used reported adult values for a clinically meaningful difference in ACT score (3-points).(17, 18) Using two-tailed Student’s t-test, a sample of 90 children (45 in each arm) will detect a difference in C-ACT of 3 points; an effect size of 0.6 given SD~5, with 80% power, at a 5% significance level. We aim to recruit 120 children, to allow for loss-to-follow-up and technical difficulty with spirometry and sputum induction. If our assumptions relating to baseline mean asthma control score (and standard deviation), retention rates and feasibility of study procedures in this age group are correct, we will have statistical power to assess the effectiveness of the intervention. If not, the data collected in this study will be useful in informing the design of a future definitive trial.

### Ethical and regulatory bodies

The study has ethical approval from the College of Medicine Research Ethics Committee in Malawi (P.04/18/2384) and LSTM Research Ethics Committee in the UK (research protocol 18-018). The study sponsor is Liverpool School of Tropical Medicine; the Clinical Research Support Unit at Malawi-Liverpool-Wellcome Trust Clinical Research Programme will provide in-country oversight.

The study protocol has been developed in accordance with Standard Protocol Items: Recommendations for Intervention Trials (SPIRIT) checklist (Additional file 1).

### Publication

Results will be published on completion in a peer-reviewed scientific journal and presented locally through the College of Medicine research dissemination conference. Results will also be shared with the participants and their families.

## Discussion

This study will provide important information on asthma-related morbidity and response to longterm treatment, in children from a low-income country. There are many challenges to providing chronic disease management in such settings, including; lack of accurate surveillance data, limited diagnostic skills, availability of treatments and patient education and adherence.(19) However, as prevention and management of non-communicable diseases move up the agenda for the poorest countries in the world, it is important to ensure that the recommended strategies are appropriate to these settings, and not merely extrapolated from high-income settings without evaluation of effectiveness.(20)

Limited evaluation of asthma guideline implementation in selected low- and middle-income countries (LMICs), has found that health care providers underutilise diagnostic tools, such as peak flow monitors, and inhaled steroid treatment, and that patient adherence to treatment is a challenge.(21, 22) Reasons for poor adherence are likely to be multifactorial, including unreliable supply of affordable medication, and misconceptions regarding effective asthma treatments.(23, 24)

In high-income settings, asthma education is considered an essential component of asthma therapy, leading to decreased emergency visits and hospital admissions (25, 26), however there are no data relating to LIC. We will explore the delivery of individualised asthma education by specifically trained, non-healthcare workers. If effective, this would be a pragmatic approach in low-income settings with shortages of medical and nursing staff. Task-shifting has been successfully employed as a cost-effective approach to deliver high-quality HIV care in Africa and there is limited evidence suggesting that this strategy may also be effective in the management of non-communicable diseases in LMIC.(27, 28) These studies have re-allocated tasks usually performed by doctors to non-physician health care workers; we will take a step further with the assessment of asthma control and health education performed by non-healthcare workers.

It is widely acknowledged that the diagnosis of “asthma” encompasses a spectrum of lower airway diseases, with diverse underlying pathophysiology (29). Sputum inflammatory phenotypes; eosinophilic, neutrophilic, mixed or paucigranulocytic, may be useful in guiding therapy, and this approach has been explored in the context of severe asthma in high-income countries (30).

However, to date there are no data regarding airway inflammatory phenotypes in wheezy children in LIC. Research from Brazil, a middle-income country, classified the majority of asthmatic children as non-atopic (31), with predominantly neutrophilic airway inflammation seen in this group, compared to eosinophilic airway inflammation seen in atopic asthmatics (32).

Children from high and low income countries are exposed to very different environments, in-utero and during early life, which may influence lung development; including maternal and infant nutrition, infections, indoor and outdoor air pollutants, and allergens. (33),(34),(35),(36),(37),(38, 39) This study will provide novel data on the clinical and inflammatory phenotypes of asthmatic children from a LIC, to inform the design of future research and the development of appropriate treatment strategies for similar resource-poor settings.

### Trial status

The current protocol is version 3.1, dated 02/04/2019. Enrolment started in September 2018 and will continue until the recruitment target is reached.

## Data Availability

Following trial completion, data will be made publicly available on an open-access repository (e.g. Mendeley Data)

**Abbreviations**
ATS: American Thoracic Society;
C-ACT: Childhood Asthma Control Test;
DSMB: Data and Safety Monitoring Board;
ERS: European Respiratory Society;
FeNO: exhaled nitric oxide;
FEV_1_: Forced Expiratory Volume in 1 second;
FVC: Forced Vital Capacity;
HFA-BDP: beclomethasone diproprionate;
ICS: inhaled corticosteroid;
LIC: low-income country;
LMIC: low- and middle-income country;
ppb: parts per billion;
QECH: Queen Elizabeth Central Hospital;
SABA: short acting β_2_ agonist.

## Declarations

## Ethics approval and consent to participate

The study has ethical approval from the College of Medicine Research Ethics Committee in Malawi (P.04/18/2384) and LSTM Research Ethics Committee in the UK (research protocol 18-018).

## Consent for publication

Not applicable

## Author’s contributions

Study design: SR, KM, CJ

Acquisition of data: SR, JP, KJ

Supervision: KM, JG, CJ

All authors have read and approved the final manuscript.

## Acknowledgements

Not applicable

Additional files

Additional file 1: SPIRIT checklist

Additional file 2: Study protocol

